# Changing contact patterns in Newfoundland and Labrador, Canada in response to public health measures during the COVID-19 pandemic

**DOI:** 10.1101/2025.09.16.25335524

**Authors:** Renny Doig, Amy Hurford, Suzette Spurrell, Andrea Morrissey, Liangliang Wang, Caroline Colijn

## Abstract

The provincial government of Newfoundland and Labrador, Canada implemented a contact tracing program as part of a containment strategy during the COVID-19 pandemic. A high proportion of cases were detected and contact traced, and our analysis provides insights into secondary case distributions and contact patterns in Newfoundland and Labrador. We used a heuristic approximation of secondary cases to account for ambiguities in who infected whom. These approximate values provide an empirical distribution of secondary cases. These distributions are compared against the stringency of public health measures. Additionally, we visualised age- and contact-based patterns and compared these patterns with respect to stringency. The maximum number of contacts traced per week was 4,645 and the mean number of contacts traced per case was 12.5. Approximate 95% CIs of the effective reproduction number under Alert levels 2-4 were (1.02,1.21), (0.99,1.39), (0.84,1.06), and (1.20,1.47). We find that this level of contact tracing was sufficient, in combination with other public health interventions, to contain pandemic SARS-CoV-2 spread in Newfoundland and Labrador prior to the establishment of the Omicron variant. Understanding age-based contact patterns is necessary to describe disease spread and the risk of severe outcomes. A successful containment strategy requires that contact tracing capacity is not exceeded, making it necessary to understand the behaviour of high-contact individuals.

## Introduction

The province of Newfoundland and Labrador (NL), Canada, announced its first COVID-19 case on March 14, 2020. In response to this the provincial government implemented a containment strategy to address the risk posed by the pandemic [1, p11-12]. The objective of a containment strategy is to prevent the spread of a disease by minimizing the risk of transmission. Containment strategies consist of several policies that test, trace, and isolate infectious individuals, that restrict both the importation of new cases and infection spread, and that are intended to interact synergistically. These strategies are distinguished from elimination and eradication strategies, which seek to reduce transmission to zero, and mitigation strategies which aim to reduce overall impact while accepting some community transmission [2–4]. Containment strategies are typically only viable during stages of a pandemic with low case counts and limited community spread. This was the case in NL until the Omicron variant of the virus was detected on December 15, 2021 [1, p59], at which point the existing containment strategies were deemed to no longer be effective. By early January 2022 the provincial government had switched to a mitigation strategy [1, p61].

Contact tracing involves identifying individuals that were potentially exposed by an infectious individual and is an important part of a containment strategy.

Contact tracing allows for the detection of asymptomatic and pre-symptomatic individuals, and the potential isolation of infectious individuals before they can generate secondary cases. The effectiveness of contact tracing, as a containment measure, often depends on both the number of contacts being traced and the proportion of cases that are successfully identified, and as capacity is reached, containment no longer becomes feasible [5–7]. Other public health measures that limit the number of people contacted by an infectious individual, or that prevent infection spread by other means, have a role in determining the efficacy of a contact tracing program, and in protecting contact tracing capacity [7–8].

In NL, contact tracing began with the first reported case on March 14, 2020 and consistent contact tracing was performed until January 4, 2022 [9], although data continued to be collected until June 2, 2022 in the pursuance of a mitigation strategy. After the public health emergency was declared on March 18, 2020 [1, p10], the NL government issued changes to public health measures via Special Measures Orders (SMOs), where these measures may have contributed to contact tracing efficacy and protecting contact tracing capacity as part of a containment strategy. The initial response to the COVID-19 pandemic involved moving classes online for grades kindergarten to twelve, travel measures, closing recreational facilities, imposing limitations on indoor gatherings, and physical distancing recommendations [1, p14]. Subsequent SMOs relaxed or further restricted these measures in response to case counts and, later, vaccination rates.

As of April 30, 2020, the NL government introduced the Alert system, which bundled together many policies affecting local transmission. Alert level 5 was the strictest set of policies and consisted of small gathering limits and closures of many activities and non-essential businesses. Alert level 2 was the most relaxed set of policies that was implemented; it permitted larger gatherings and businesses to operate conditional on physical distancing and masking guidelines [1, p17-19]. Alert level 1 was a return to a “new normal” and was never implemented, as instead the Public Health Emergency was repealed on March 14, 2022 [1, p71].

The Alert level system did not encompass all of the component policies of NLs containment strategy. Travel measures, suspension of in-person kindergarten to grade twelve school, and mandates on indoor masking were issued in separate SMOs and were not directly linked to the Alert levels. Additionally, while gathering limits were not fixed for each Alert level, the stringency of the gathering limits tended to correspond to the stringency of the Alert level.

The implementation of stricter public health measures may change age-group contact patterns, and other aspects of contact assortativity. Characterizing such changes helps to accurately assess the risk of severe infection, which is greatest for older individuals. Assortativity patterns, such as individuals with a high number of contacts also contacting other individuals with a large number of contacts, affect the risk of an outbreak that grows quickly due to consecutive super-spreading events. Understanding the risk of quickly-growing large outbreaks is necessary to understand whether contact tracing capacity is likely to be exceeded.

We use the data collected by NL through their contact tracing program to understand the efficacy of the containment strategy implemented by the NL government during the COVID-19 pandemic. We report contacts traced per week and contacts per case for different stringencies of public health measures (Alert levels) to understand the capacity of NL’s contact tracing program. We report the number of travel-related and community cases, and estimate distributions of secondary infections and cluster sizes to evaluate the efficacy of NL’s containment strategy. We visualize contact patterns for age-groups and assortativity based on the number of contacts to better understand the outbreaks that occurred in the NL population.

## Methods

### Data collection

The contact tracing program in NL was carried out by a core team of 15 public health officials. When an eligible individual was tested, a public health official would call the individual to inform them of their test result. If the test result was positive, a list of recent contacts was obtained, contacts being defined as any individuals who the case had close contact with within 72 hours of the case’s episode date (episode date being date of symptom onset if the case was symptomatic, or date of specimen collection if the case was asymptomatic) [10].

Later in the pandemic, individuals could learn of their test result by checking an online system. When results were communicated online, public health officials monitored the test results database for the purpose of initiating contact tracing as quickly as possible when positive cases were reported. Contacts of a case were instructed by either a letter and/or a telephone call to test and isolate for 14 days. If the individual tested positive, then the duration of the isolation period would be reduced to 10 days from symptom onset, or date of positive test for those asymptomatic. Contacts of a case were usually contacted by the contact tracers within 24 hours. During periods of many cases, additional staff and third parties were contracted to support the core team of contact tracers. During the study period, all contacts were notified as described, such that contact tracing was conducted consistently. On December 15, 2021, it was stated by the Chief Medical Officer of Health that due to the short generation time of the Omicron variant, contact tracing could no longer be completed with the same efficiency as had occurred prior, and this is how we defined the end of our study period.

For the duration of the study period, symptomatic individuals in NL could seek testing if they met testing criteria at that moment in time. Testing criteria shifted throughout the study period; at times one symptom could screen clients in for a COVID-19 test while at other times two symptoms were required, one of which had to be fever or cough. Furthermore, some asymptomatic individuals were also required to complete tests: rotational workers, with some modifications to the corresponding self-isolation rules [1, p24-25], close contacts of a confirmed case, and individuals that were asked to complete testing due to publicly issued exposure notifications corresponding to a venue, flight, or ferry. There were also testing requirements for entering some long-term care facilities, personal care homes, assisted living facilities [1, p17], and health care facilities.

For positive cases, the source is recorded as being related to travel, a close contact of a traveler, or a local source. Additionally, geographic information is recorded in the form of the Forward Sortation Area (FSA) of the individual. Due to the relatively small amount of local spread during the focal period of this study, the contact tracing networks observed are assumed to be mostly complete.

### Data preparation and cleaning

We organized the contact tracing data into distinct contact networks, which are defined in this paper as a collection of individuals who are connected by at least one contact event. A contact event occurs between individuals A and B if A lists B as a close contact or vice versa. The contact event alone does not indicate who infected whom . Compiling connected contact events into clusters results in two categories of cases: those that are only travel-related and those that eventually result in local spread. If a contact network contains only individuals who are recorded as having a history of travel or as being a close contact of a traveler, then all individuals in the cluster are labeled “travel-related.” Otherwise, a network has at least one instance of a contact event which constitutes local spread. Since the policies of interest in this work are the Alert level measures, which target local spread, travel-related networks are removed for the analysis. Within each contact network is a case cluster, which contains all of the individuals in the network that tested positive.

The Alert level that was in place at the time and location of each contact event is not recorded during the contact tracing process. To compare the structure of the contact networks between Alert levels, each contact event was linked to an Alert level based on the FSA in which the contact happened. The Newfoundland and Labrador, Department of Health and Community Services Public Service Advisories placed certain geographic regions under particular Alert levels at different times. Each geographic region was assigned a FSA that most closely matched it. The Alert level for each contact event was the Alert level assigned to the FSA at the time the contact was reported to have occurred. It is noted that since SMOs affecting the Alert level were not issued based on FSA there may be minor discrepancies between the assigned Alert level and the true Alert level for some contact events.

During the data cleaning process, any cases that occurred outside of the focal period were removed. Cases that were travel-related were removed as well as any entries that had missing or invalid FSA information.

### Modeling the number of secondary infections

The effective reproduction number, R_t_, is the average number of secondary infections resulting from one index case in a partially susceptible population at time *t*. When complete contact tracing data is available, R_t_ can be estimated directly as secondary case counts are available for each case. In cases where available data is limited to time series of reported cases or symptom onset, direct estimation of R_t_ is not possible and some model structure needs to be applied to leverage some information about secondary case distributions from the data. Mechanistic models of transmission dynamics are common [11], as are models that have a more limited structure, assuming only a probability distribution over the interval of time between cases [12,13]. In both cases, the lack of direct observation of contact events between individuals necessitates some additional assumptions/structure. The contact tracing data used in this study is partially complete in that we observe contact events, but not directly who infected whom. As such, we adopt a heuristic approach, described below, which makes use of the partial information available without having to rely on a structured model requiring additional parameters to be estimated or assumptions to be made.

The number of secondary cases generated by a particular index case, *i*, is denoted *I_i_*. To account for heterogeneity in infectiousness, *I_i_* is often modeled as a negative binomial random variable with mean R_t_ and precision parameter *k* [14]. Under this parameterization of the negative binomial distribution the number of secondary cases has variance 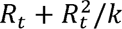. As the estimated value of the precision parameter for our data was large relative to 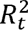, we also consider a Poisson distribution with mean R_t_, as it is the limiting distribution of a negative binomial as the precision parameter gets very large.

Observing the number of secondary cases requires knowledge of the infector and the infectee in each contact event. As an example of a contact network for which the infector and infectee labels can be assigned, consider a contact network containing two individuals, A and B, such that A travels and tests positive, then lists B as a contact who then tests positive without coming into contact with anyone else. Clusters including multiple travellers or more interconnected individuals do not admit a clear infector/infectee label for each individual, therefore direct observations of *I_i_* are not available. To compensate for this, an approximate number of secondary cases, ̃*I_t_*, is computed. Each individual in the dataset will either be an index case or they will have at least one other individual listing them as a contact. The number of cases listing an individual as a contact is the number of potential infectors that the individual has. Take three individuals, A, B, and C, who have n_A_, n_B_, and n_C_ potential infectors, respectively. Now consider a fourth individual, D, who is a potential infector of all of A, B, and C. Individual D constitutes 1/n_A_-th of A’s potential infectors, and similarly for B and C. So, to approximate the number of secondary cases attributable to D, we define 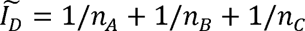. These ̃*I_t_*’s are then modeled using a continuous version of the above negative binomial and Poisson distributions, swapping any factorials in the probability mass function for the appropriate gamma function. This weighting procedure is intended to be a heuristic approach, used so that a parametric estimate of the effective reproduction number can be obtained. A point estimate and an approximate 95% confidence interval about *R_t_* is obtained via maximum likelihood estimation.

### Visualizing contact patterns

Two types of plots are used to visualize the contact structure within the clusters. The first is an age-stratified contact matrix. Each individual is placed into an age group: [0,5), [5,18), [18,30), [30,40), [40,50), [50,60), [60,70), and 70+. The age groups were chosen to reflect pre-school-age ([0,5)), school-age ([5,18)), and then 10-year bins after the age of 18. The grouping together of school-age individuals is supported by a study conducted in Tianjin, China [15], as well as contact matrices estimated in Canada [16] and the United Kingdom [17], which show very similar contact patterns for these ages. The number of contacts between each age group are counted and plotted as a heat map that shows the total number of interactions between age groups within the observed contact networks. Counts below five are shown as being exactly five to preserve anonymity. The second type of plot shows the relationship between the number of contacts reported by each pair of cases connected by a contact event. For each pair of connected individuals, the number of contacts reported by each was computed and plotted as a point, with the initial case on the horizontal axis and the contacted case on the vertical axis. Rather than plotting the individual points, this pattern is shown by applying 2-dimensional kernel smoothing to these points and plotting the contours of the smoothed density. The darker contours represent regions of case/contact pairs where there were a relatively higher number of contact events with that pairing of reported cases.

Both visualizations are computed for all data in the focal period disaggregated into the Alert level during which the contact event took place. In addition to these aggregate plots, the same plots are computed for two individual clusters. The two clusters that are selected contain the most contact events. All analysis is done using the R programming language [18]; the ggplot2 package [19] is used to generate all figures.

## Results

An overview of the history of the disease between the period of March 14, 2020 and December 15, 2021 is given in Figure 1, which shows weekly aggregate cases in the province. Reported cases are separated into those that are travel-related and those constituting local spread; local spread is then further separated into the Alert level in effect when the contact event occurred. Changes to provincial masking mandates are shown separately from the Alert levels, as these policies were kept separate. The period between March 14, 2020 and April 30, 2020 is shaded grey and corresponds to the period prior to the Alert level system; bars in this period are coloured black because there is no Alert level to which they correspond. After an SMO that placed the province at Alert level 5, subsequent relaxations brought the Alert level province-wide down to level 2 by July 2020 when there were no instances of community spread. In late 2020 and early 2021, when the entire province was at Alert level 2 and there was an indoor mask mandate in effect, there were several weeks with reported cases. However, most of these cases remained travel-related and did not result in community spread. In February 2021, when a large amount of community spread was detected on the Avalon peninsula of Newfoundland, the entire province moved to Alert level 5. The implementation of measures at the community-level, rather than province-wide, started shortly after this on February 27, 2021; the province continued to manage community-level policy changes for the remainder of our focal period. The case peak in early summer 2021 was largely restricted to travel-related cases, while the peak in late summer/early fall, which began shortly after the mask mandate had been lifted, was dominated by community spread and a mask mandate was reissued a few weeks later. Protecting contact tracing capacity was one stated rationale for re-issuing the mandatory mask requirement, and at this time more than 1,000 contacts were being traced per week (Figure 1, top panel). At two points within the focal period, February 2021 and September 17, 2021 [1, p49; 17], the NL government reported a strain on contact tracing capacity. The core contact-tracing team in eastern Newfoundland consisted of fifteen public health officials [20]. Figure 1 shows the weekly total of contacts traced during the period (top panel). Peaks in the number of contacts traced tend to correspond to peaks in reported cases, but on a larger scale.

**Fig. 1.**
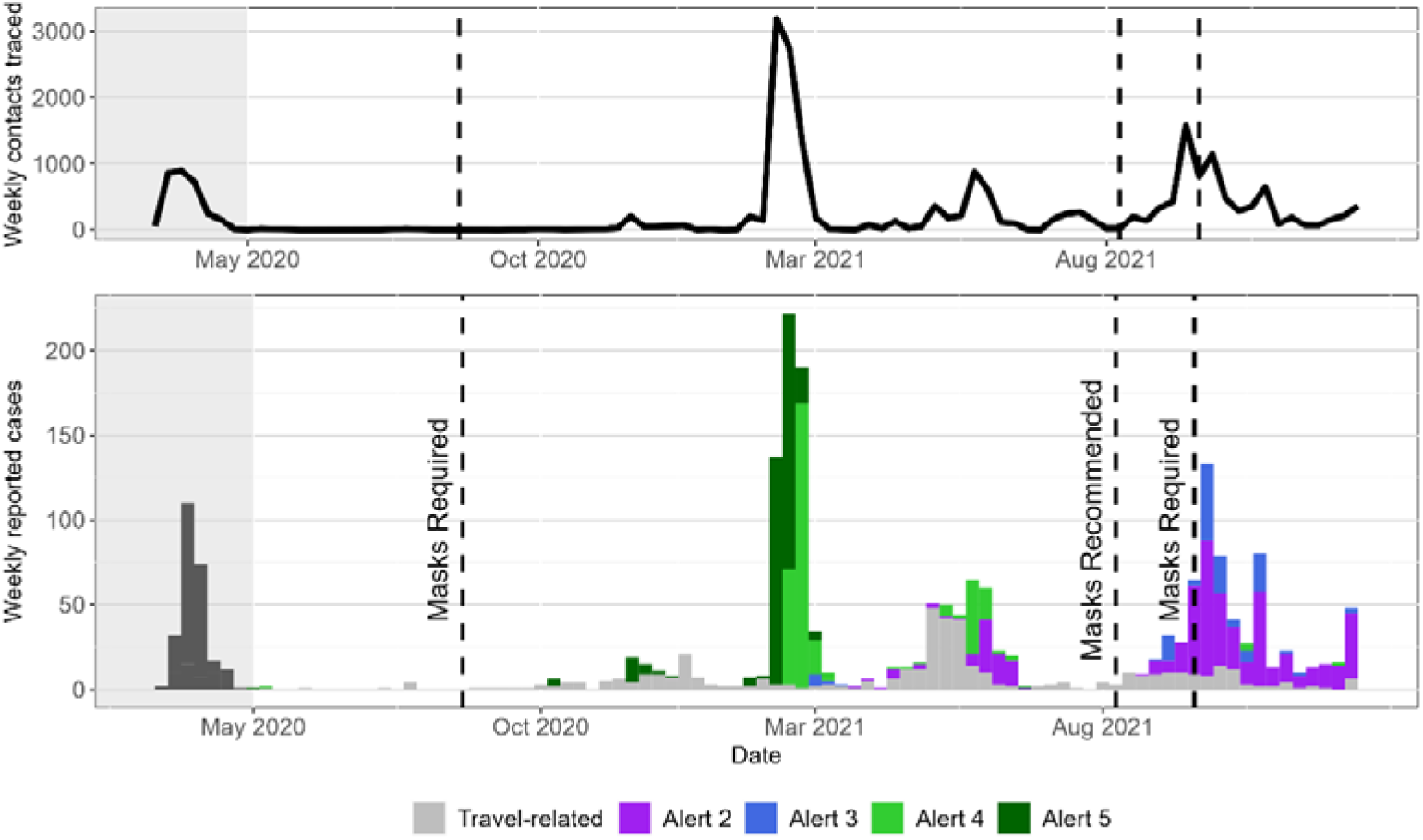
Weekly cases (bottom) and contacts traced (top) in Newfoundland and Labrador until December 15, 2021 with colours indicating if the case is travel-related or if not, and which Alert level it occurred under. Vertical dashed lines indicate province-wide mandates on masking indoors.

Between March 14, 2020 and December 15, 2021, 22,626 contacts were traced. Of these contacts, 2,111 were cases, 1,522 of which were related to local spread. Analyses of the Alert levels focused on data occurring after April 30, 2020, removing 203 cases. A further 7 cases were removed for having incorrect or missing FSA data. A numerical summary of the contact tracing efforts is supplied in Table 1. The table shows the total number of contacts traced, the total number of cases reported, the maximum number of contacts traced in one week, and the average number of contacts traced per person; these results are shown for the entire focal period as well as a breakdown by Alert level. Of the 1,522 cases, 1,299 (85.3%) reported symptoms, 71 (4.66%) were hospitalized, and 17 (1.12%) died.

**Table 1.** Numerical summaries of contact tracing during focal period.

|  | Overall | Alert 2 | Alert 3 | Alert 4 | Alert 5 |
| --- | --- | --- | --- | --- | --- |
| Contacts traced | 19269 | 5740 | 1484 | 3199 | 6004 |
| Cases reported | 1522 | 484 | 137 | 349 | 342 |
| Max. contacts traced in a week | 4645 | 1217 | 406 | 996 | 4645 |
| Mean contacts traced per person | 12.5 | 11.9 | 10.8 | 9.17 | 17.6 |

### Observed transmission clusters

There were 166 distinct clusters containing more than one person during the focal period. The largest cluster contained 276 cases. The median cluster size was 3, with 90% of the clusters containing at most 11 cases. The distribution of cluster sizes across the whole focal period is illustrated in Figure 2. Both plots show a distribution of cluster sizes concentrated on small numbers of cases with a long tail.

**Fig. 2.**
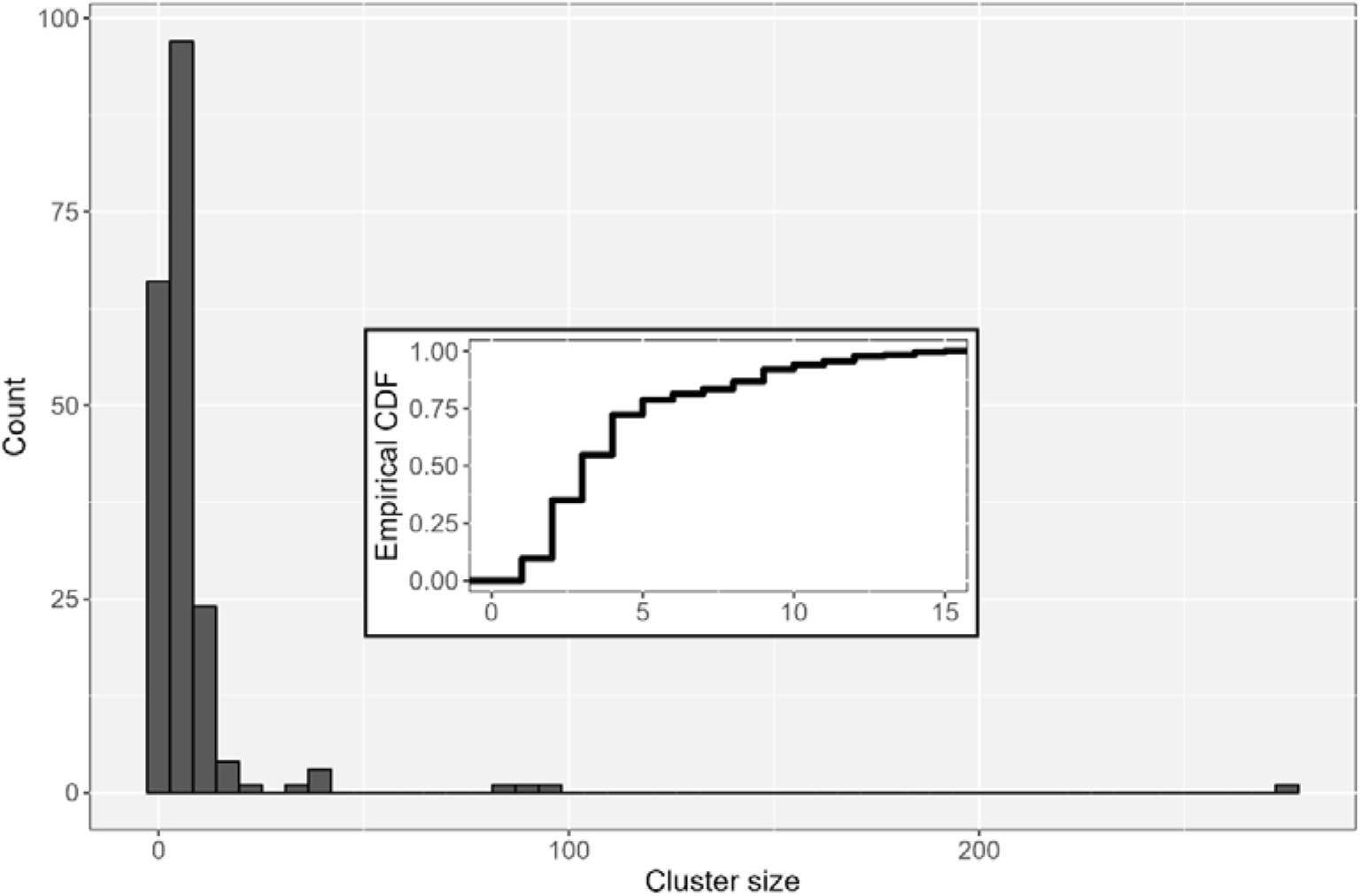
Histogram of the cluster sizes across the focal period; inset contains the corresponding empirical cumulative distribution function, truncated to 15.

The largest contact network during the focal period is shown in Figure 3. In addition to the case cluster itself, shown with the larger opaque points, Figure 3 also shows individuals who were traced but did not test positive in the smaller, translucent points. This network is linked to an outbreak in eastern Newfoundland between February and April, 2021. Of the contact events that comprise the network, 38.7% occurred under Alert level 5, 60.8% occurred under Alert level 4, and 0.5% occurred under Alert level 2. At the centre of the cluster most of the contacts fall within the 5-17 age group. There are also several contacts that are in the 30-59 age groups. The smaller points indicate contact events which did not produce secondary cases either because transmission did not occur or because contact tracing contained the infection.

**Fig. 3.**
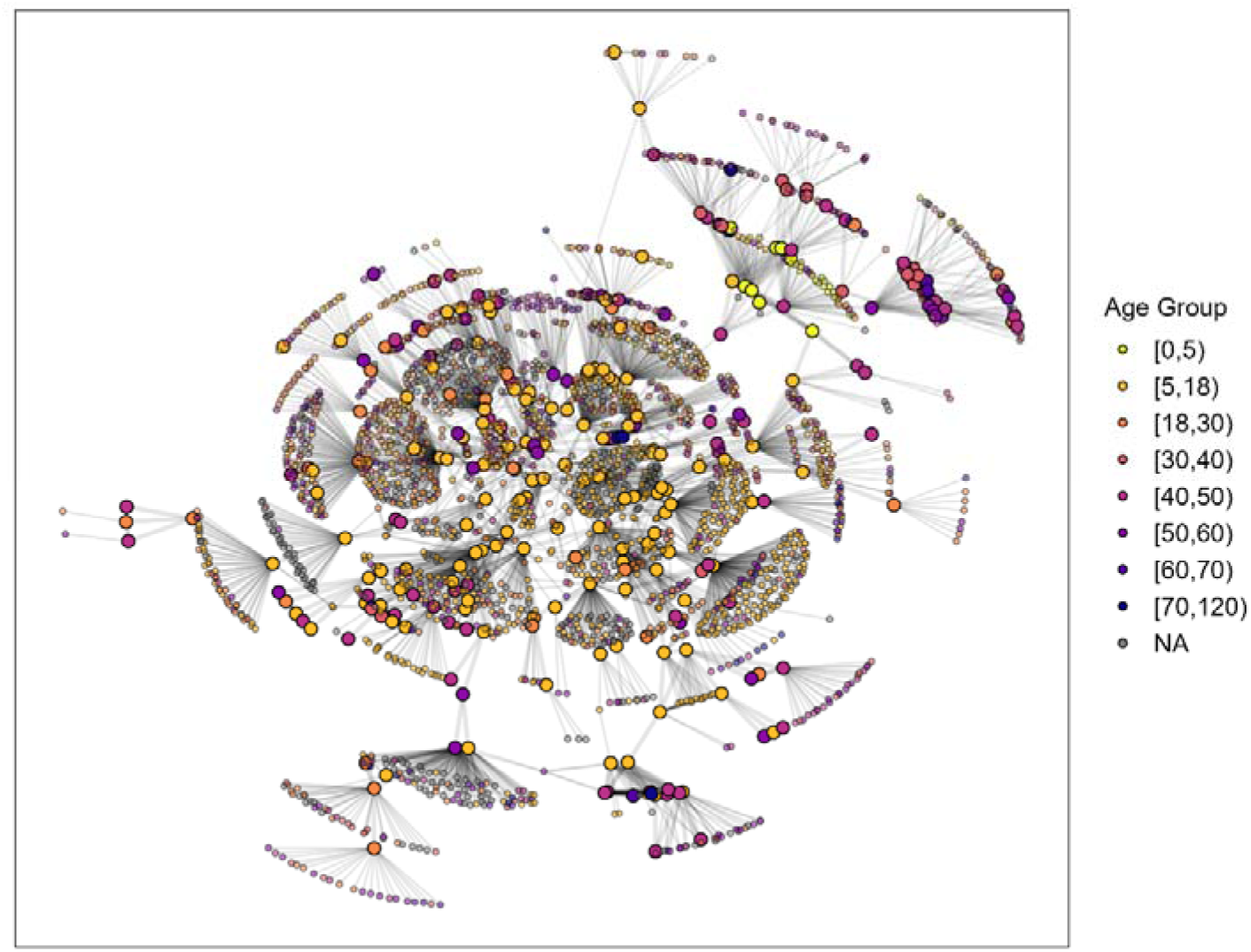
Graph of the largest contact network in the dataset. Large, opaque points are the cases which comprise the cluster itself and the smaller, translucent points are contacts which were traced but did not result in further local spread. The opacity of the edges corresponds to the number of subsequent contacts resulting from that infection.

### Effective reproduction number

The smoothed empirical distributions of secondary cases are visualized in Figure 4 for the period from October 2020 until the end of the focal period. The values of approximated secondary infections, ̃*I_t_*, are smoothed over weekly periods and are disaggregated by Alert level. The corresponding plot of weekly cases is shown above. There are three predominant peaks visible during this period; the first occurring under Alert levels 4 and 5, the second under Alert levels 2 and 4, and the final mostly under Alert levels 2 and 3. For most weeks the distribution of secondary cases appears to be centred around 1, skewing more towards 0 as the outbreaks taper off.

**Fig. 4.**
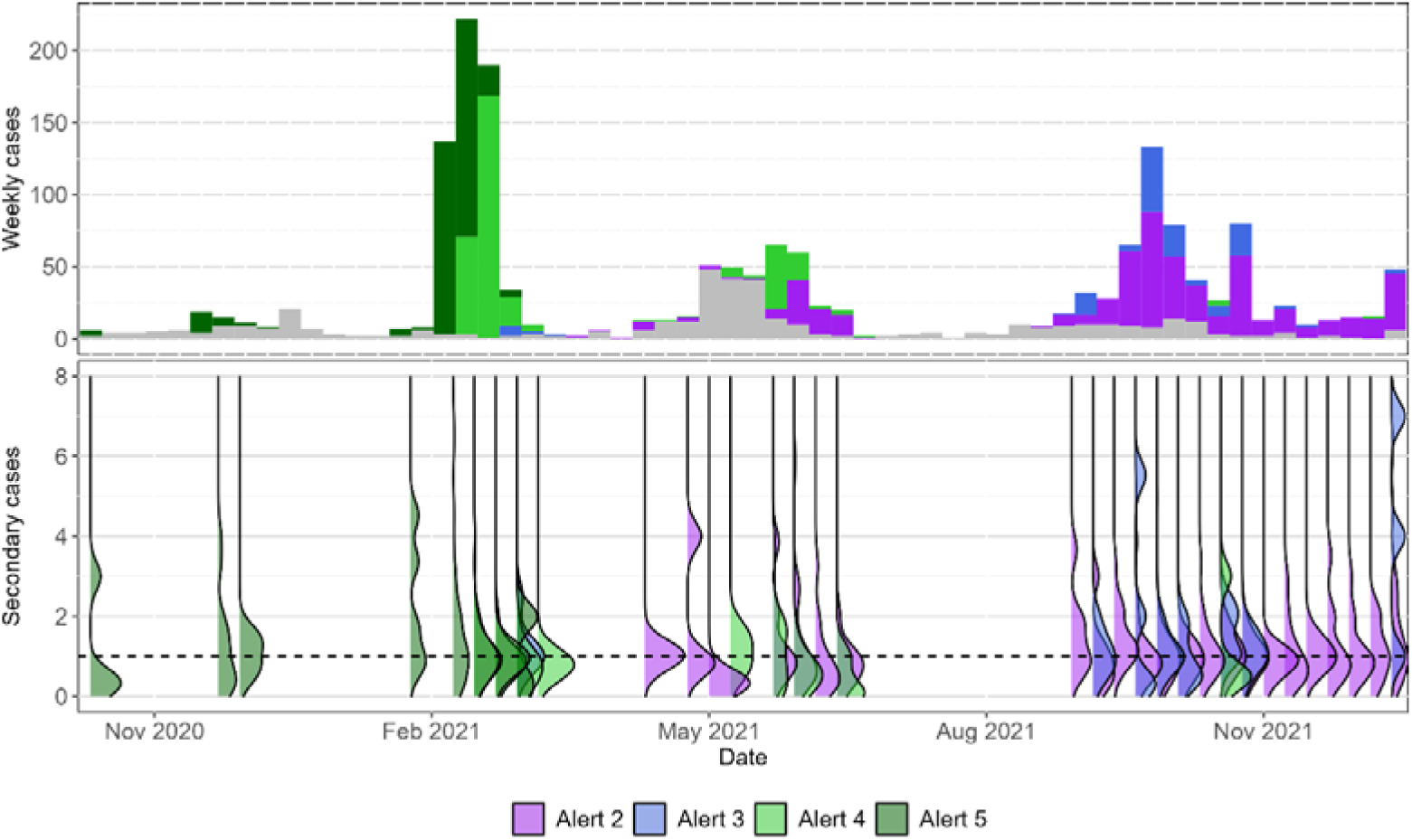
Weekly cases between October 2020 and December 15, 2021 (top) and smoothed empirical distribution of secondary cases, pooled over a weekly period, (bottom).

The secondary cases were then aggregated across the entire focal period for each Alert level; the corresponding histograms are shown in Figure 5. Both the negative binomial and Poisson distributions were fit to the data in each Alert level via maximum likelihood and compared using the Akaike Information Criterion (AIC). The fitted Poisson distribution resulted in a lower AIC for all Alert levels, so the estimates below correspond to the Poisson distribution. It is noted that the point estimates for the reproduction number were very similar under the negative binomial distribution, however the likelihood function was very insensitive to the value of the precision parameter. Maximum likelihood estimates (MLEs) for the reproduction numbers and precision parameters, as well as the AIC for each Alert level are shown in Table 2. The MLEs of *R_t_* for each Alert level are also shown in Figure 5 along with an approximate 95% confidence interval (CI). The probability mass functions, computed using the MLEs, are drawn over the histograms.

**Fig. 5.**
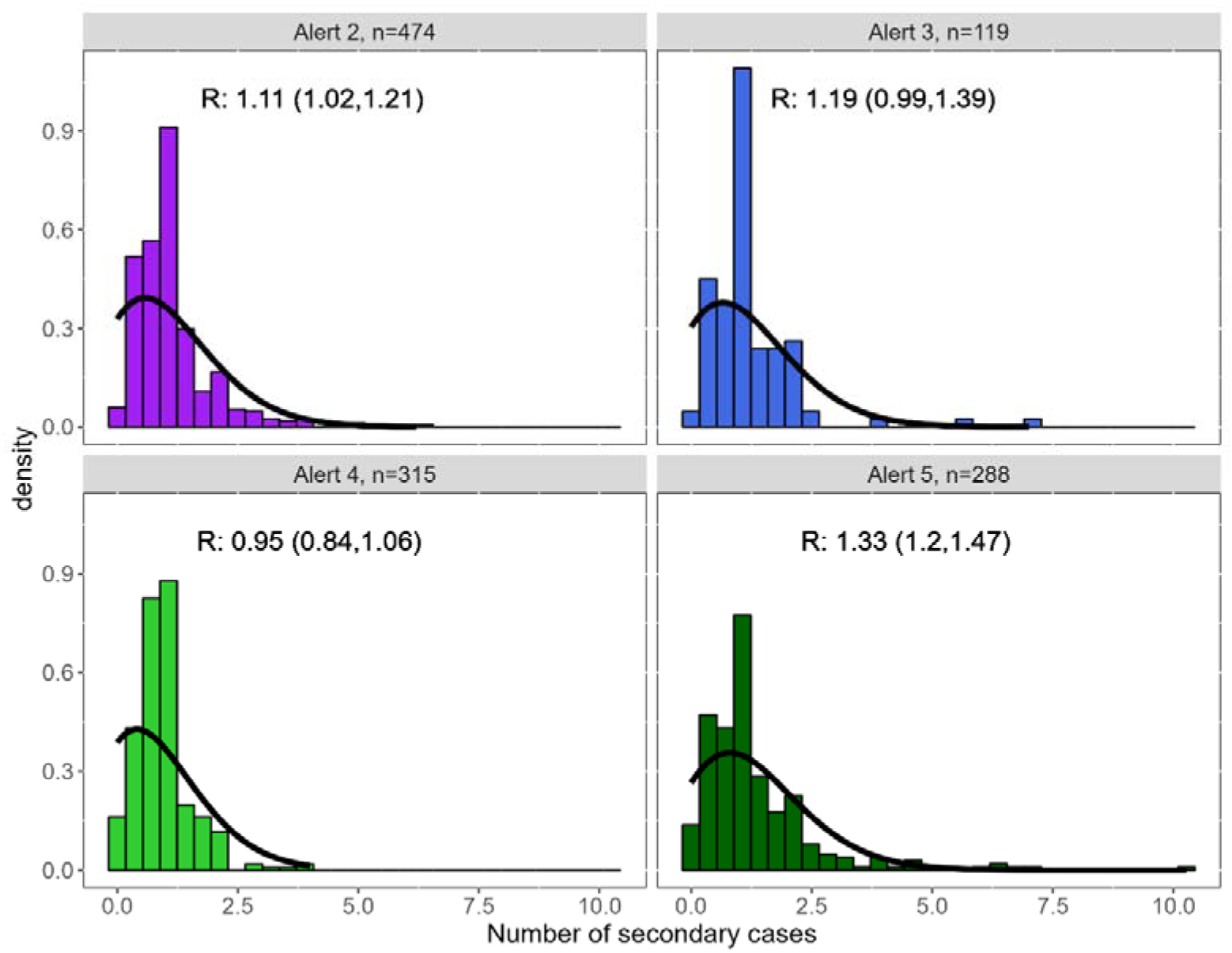
Histogram of the estimated number of secondary cases caused by each index case under each Alert level. Probability mass functions evaluated at the MLEs are shown with a solid line. Approximate 95% CIs for *R_t_* are shown in the top right corner.

**Table 2.** Point estimates and 95% CIs for *R_t_* under each Alert level.

| Alert Level | Poisson |  | Negative Binomial |  |  |
| --- | --- | --- | --- | --- | --- |
| | $\widehat{R}_t$ | AIC | $\widehat{R}_t$ | $\hat{k}$ | AIC |
| 2 | 1.11 | 1148 | 1.11 | 194.0 | 1151 |
| 3 | 1.19 | 298 | 1.19 | 58.7 | 301 |
| 4 | 0.95 | 680 | 0.95 | 107.0 | 684 |
| 5 | 1.33 | 825 | 1.33 | 46.8 | 826 |

### Contact patterns

The visualizations of contact patterns within the clusters are shown for all clusters within each Alert level as well as for the two largest clusters observed after April 30, 2020, with respect to the number of contact events. The first cluster is the largest observed cluster with 605 contact events and 276 cases, occurring in the former eastern health region (Newfoundland) between February and April, 2021 and the second largest cluster contained 261 contact events and 91 cases and occurred in the former central health region (Newfoundland) between May and June, 2021. Of the contact events that comprise the central cluster, 65.9% of the contacts occurred under Alert level 4 and 34.1% occurred under Alert level 2.

Subsequent references to these clusters will refer to the region in which they occurred. The four regional health authorities of Newfoundland and Labrador were amalgamated in April 2023.

Age contact matrices are shown in Figure 6. In Alert levels 2 and 3 the contacts are fairly dispersed across age groups and show minimal clustering around the diagonal. Alert level 4 shows a relatively large proportion of contacts occurring within and between the 5-59 age groups. Alert level 5 shows strong interaction between the 5-17 and 40-49 age groups. All four plots show the highest number of contacts occurring in the 5-17 age group.

The eastern health cluster shows predominantly cases in the 5-17 and 40-49 age groups with interaction between these age groups. Other ages are present, in particular a collection of contacts in the 30-59 age groups. There are some similarities between the patterns present in the eastern health cluster and those for Alert level 5. Given the relative scales of the two plots, an explanation for this could be that the eastern cluster is driving the pattern for the entire Alert level. The plot for the central health cluster shows an arrow-shaped pattern with a high concentration in the [18,30) age group as well as interactions between this group and the [30,60) age groups. Notably, there is much less interaction between the [30,60) age groups than there is between them and [18,30). There is also a more uniform pattern occurring between the [5,18) and [18,30) age groups. The pattern present for the central cluster does not bear a strong resemblance to any of the Alert level patterns. This is an example of a case where population-level trends in age assortativity for an Alert level are not reflected in individual clusters.

**Fig. 6.**
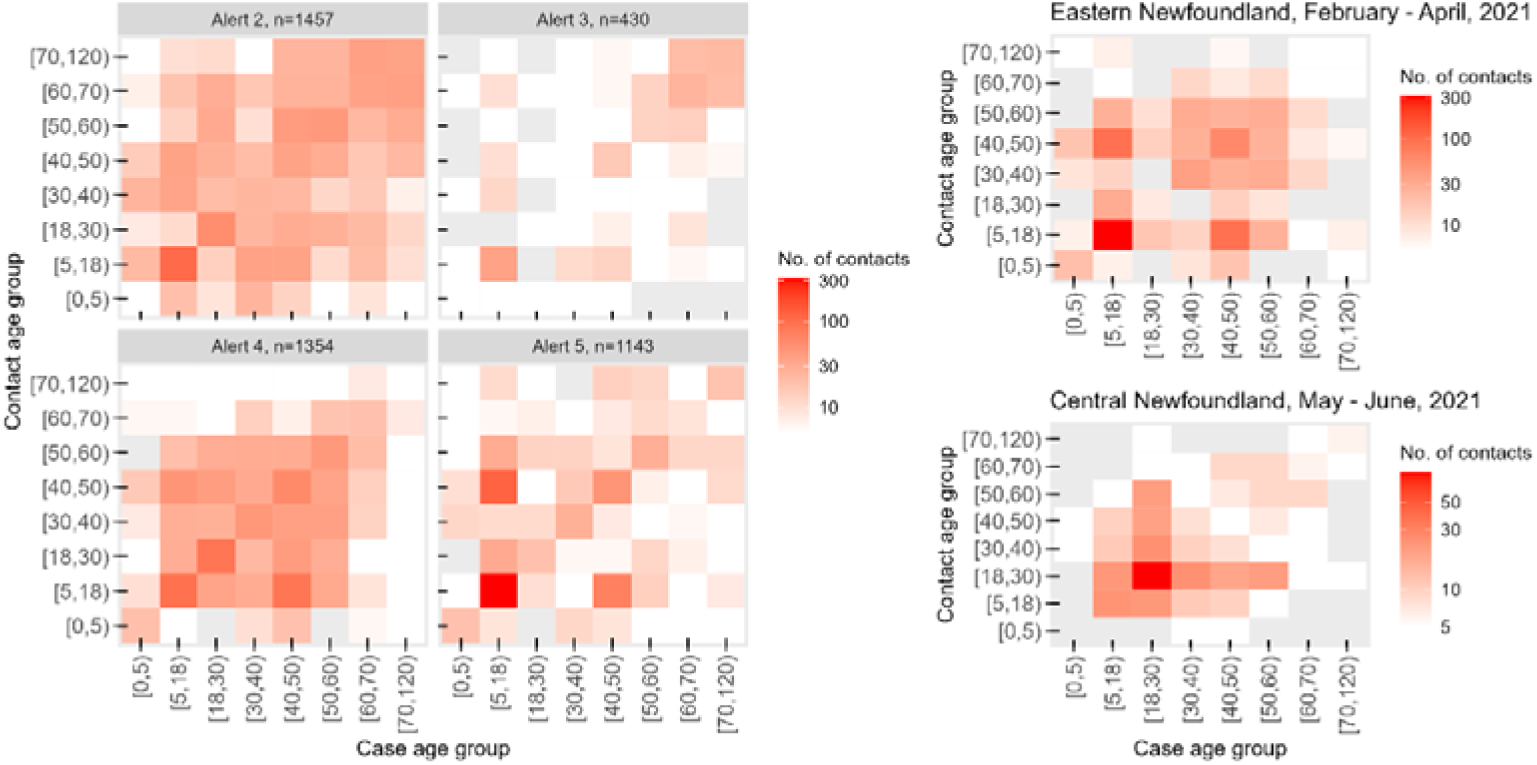
Contact matrices indicating the number of contacts that occur within and between age groups. The panel on the left shows matrices computed for each of the Alert levels separately, aggregated over all contacts occurring under that particular Alert level. On the right are the same matrices computed from the eastern and central clusters.

The density plots of the high-contact/low-contact interaction patterns are shown in Figure 7. Alert levels 2 and 3 both show patterns with a single point of concentration and some dispersion away from the diagonal as the number of contacts increases. In Alert level 2 the contact frequencies are centred about the point (6,6), meaning that for many of the cases with around 6 contacts, each contact themselves produced around 6 contacts. Alert level 3 shows a similar pattern, but more dispersed and slightly more skewed towards high-contacts/high-contact pairs. Alert levels 4 and 5 differ in that they have multiple points of concentration and less consistent dispersion. Alert level 4 has one point of concentration around (5,5), but with another localized concentration at a high-contact/high-contact point on the diagonal. The number of contacts per-person in Alert level 5 is quite dispersed. In all cases, most individuals have fewer than 20 contacts. The eastern cluster shows two concentrations: one in the low-contact corner and another at a higher number of contacts. This plot shows a clear interaction between high-contact and low-contact individuals (paler areas off the diagonal). The central cluster has three areas of concentration. The highest concentration is around 40 contacts per person, with a lower concentration at the low-contact/low-contact corner of the plot, and low-contact/high-contact mode located symmetrically about the diagonal. While the central cluster had few cases, it tends to show a much higher number of contacts per person.

**Fig. 7.**
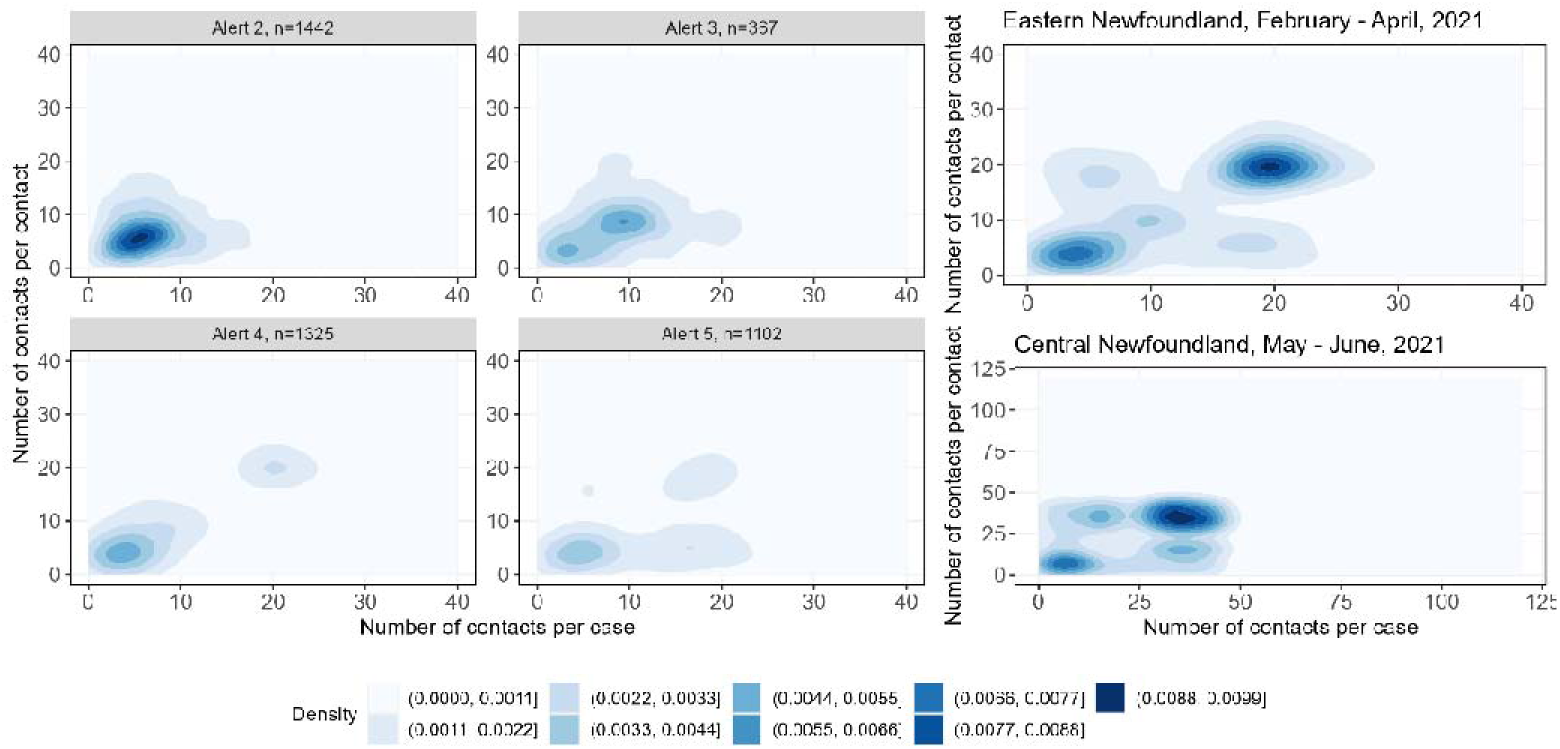
Joint distribution over the number of contacts in each case/contact pair; each region (i.e. each colour) contains all pairs with similar estimated density values, as indicated in the quantile legend. The left panels show the contours constructed from data aggregated for each Alert level. The panels on the right show the contours for the eastern cluster and the central cluster.

## Discussion

Between March 20, 2020 and December 15, 2021, the NL government implemented a contact tracing program as part of a COVID-19 containment strategy. The data provides information about secondary cases and contact patterns in the population. Secondary case distributions can indicate how effective policy measures are at suppressing local spread. Analyzing contact patterns under varying levels of policy stringency can either validate existing assumptions of contact homogeneity or provide a basis for more accurate models of disease transmission. NL’s public health measures until the establishment of the Omicron variant were effective at containing SARS-CoV-2 spread (Figures 1 and 4), and the contact tracing effort to achieve these results is summarized in Figure 1 and Table 1.

The distributions of secondary cases (Figures 4 and 5) and the estimates of *R_t_* (Figure 5) offer insight into the efficacy of the Alert level system. The shapes of the secondary case distributions in Figure 4 are skewed above 1 during periods of increasing incidence and skewed below 1 as peaks taper off. The estimates of *R_t_* for each Alert level are mostly close to 1 with 95% lower confidence bounds of 1.02, 0.99, 0.84, and 1.20. This is consistent with local spread being contained until late December 2021. The higher value for Alert level 5 could be a consequence of low compliance with public health measures prior to the February 2021 outbreak in the eastern health region [1, p28], and delays between identifying the outbreak and the implementation of Alert level policies. The results here should be evaluated while keeping in mind that the heuristic nature of the estimation procedure precludes rigorous statistical interpretation. Furthermore, Figure 1 shows that the Alert levels are not distributed uniformly across the focal period; a consequence of this is that factors such as variant or vaccination rates could be affecting our estimates of *R_t_*.

Statistical evidence supporting the use of a Poisson distribution to model secondary cases over the negative binomial distribution typically coincides with low heterogeneity in transmission, which has not been characteristic of COVID-19 [21,22], though high uncertainty in the precision parameter has been observed in settings with very stringent control measures [23]. Additionally, it has been observed that imposing control measures can reduce overdispersion [24].

Therefore, one plausible explanation for this deviation from the existing COVID-19 literature, with respect to the secondary case distribution, is that control measures in place, even those with lower stringency, have reduced the occurrence of superspreader events and resulted in a more homogenous pattern of infectivity for our dataset.

The age contact matrices shown in Figure 6 show that the 5-17 age group had the most contacts and many of those contacts occur between individuals in that age group. Additionally, the matrix for the eastern cluster showed interaction between this age group and the 40-49 age group. These observations could be supported by transmission between high school students which either is preceded by or proceeded from transmission between students and their parents/guardians [10,17]. These patterns can also be seen in Figure 3, with many individuals in the [5,18) age group at the centre of the cluster and smaller clumps of individuals in the [40,50) age group. The pattern present in the central cluster suggests a form of transmission facilitated by the [18,30) age group with minimal interaction between the older affected age groups. This represents a very different form of interaction than the former cluster. Overall, trends in interaction between age groups are most pronounced under the most stringent set of public health measures with trends becoming less apparent as stringency decreases. Comparing the contact matrices of the eastern and central clusters to the Alert-level clusters illustrates how contact patterns aggregated across multiple clusters may not be representative of patterns within particular clusters. This suggests that during the early stages of an outbreak, models which reflect contact patterns specific to the outbreak setting may be a more reliable basis for local policy decisions than aggregate models.

Age contact matrices have been estimated for all of Canada during the period May 2020 to December 2020 [16]. These showed a relatively high number of contacts occurring within the same age group with the most contacts occurring within the 5-17 age group. These contact matrices also show older age groups have more between-age interactions with a higher number of interactions among adults aged 18-60 and a lower number for those of retirement age. These patterns are similar to, but less pronounced than, those found in the POLYMOD study [17]. These Canada-wide patterns are most similar to those observed in NL during Alert level 2 (Figure 6). The block-like behaviour of the 18-60 age groups is also found in NL during Alert level 4. However, both the patterns observed during Alert level 5 and those observed in the two selected clusters have no clear resemblance to either the Canada-wide study or the POLYMOD study. This underlines the importance of understanding local contact patterns when modelling the spread of an infectious disease in small populations, or during periods of low incidence.

The interaction between low-contact and high-contact individuals was shown in Figure 7. These plots provide some information about how the distribution of the number of contacts per case varies across the population. A containment strategy requires that contact tracing capacity not be exceeded, as such it is valuable to investigate potential pressure on contact tracing demand arising from high-contact individuals. Alert levels 2 and 3 both show a pattern of high concentration in one area of the plot with greater dispersion as the number of contacts per case increases. This suggests that given an individual has a high number of contacts, there will be higher variance in the number of secondary contacts associated with them, compared to an individual with a small number of contacts. This pattern could be explained by each individual’s number of contacts being drawn from a common distribution with one mode and a skew to the right. On the other hand, Alert levels 4 and 5 exhibit less consistent behaviour with multiple points of concentration. This could be explained by the more stringent measures both limiting gathering sizes (low-contact modes) while still permitting some essential, high-contact jobs to be performed (higher-contact modes). That we observe a concentration of low-contact cases in each Alert level is another indication that the Alert level system was effective in limiting community spread. The results for the two example clusters are less neatly concentrated patterns. In the eastern cluster there appear to be two main concentrations of contacts per case, with an additional, smaller concentration reflected off the diagonal. Comparing this plot to the corresponding age contact matrix suggests a potential relationship between the age of a case and the distribution of the number of contacts per case. The patterns of the central cluster are quite similar to the eastern cluster, although at a slightly greater magnitude. Contrasting this with the age-based contact patterns, which were different between the two example clusters, suggests that whatever demographic or environmental variables drive one form of contact patterns do not affect the other in the same way. The difference in magnitude between the eastern and central clusters also provides additional support for the efficacy of the Alert system since many of the individuals in the central network had a high number of contacts, even though the resulting cluster had fewer cases than the eastern cluster.

Many infectious disease transmission models rely on strong assumptions about the contact patterns, e.g. homogeneous mixing, or on contact patterns which are estimated *a priori*. The observed contact patterns do not support a model of homogeneously mixing populations and supports previously made observations [15] that contact patterns change considerably in response to public health measures. This degree of change, which goes well beyond simply having overall contact volumes rise and fall with control measures, should raise notes of caution for infectious disease transmission modelling that relies on network and contact structure. Contact tracing provides a double benefit to infectious disease modellers and public health officials. In addition to its role in effective containment strategies, clear contact tracing data can provide modellers with information about person-to-person contact patterns which enable more effective modelling of low-incidence disease outbreaks, such as those in small populations and during the early stages of an outbreak.

## Conclusion

Estimated effective reproduction numbers are consistent with periods of contained local spread. Further analysis needs to be done to determine the individual effects of the Alert system, vaccination rates, and variant, as well as the interactions between them. The data revealed heterogeneity in contact patterns within the population. The patterns varied between contact networks as well as between individual networks and patterns aggregated over multiple networks. These suggest that population-specific age-based contact patterns are important to modelling disease spread in small populations. Additionally, given the importance of contact tracing to a containment strategy, understanding the behaviour of high-contact individuals may be crucial to keeping cases within contact tracing capacity.

## Abbreviations

*AIC*: Akaike Information Criterion
*CI*: Confidence interval
*FSA*: Forward Sortation Area
*MLE*: Maximum likelihood estimate
*NL*: Newfoundland and Labrador
*SMO*: Special Measures Order

## Declarations

### Ethics approval and consent to participate

Ethics approval for this project was granted by the Newfoundland and Labrador Health Research Ethics Board (reference # 2021.013).

### Consent for publication

The Newfoundland and Labrador Health Research Ethics Board approved the research involving secondary use data without requiring further consent to participate from the participants (i.e., Article 5.5A of the Tri-Council Policy Statement 2).

### Availability of data and materials

Data requires ethics approval and can be requested from Newfoundland and Labrador Health Services, Digital Health, the data custodian.

## Authors’ contributions

RD performed the analysis, generated visuals, and wrote the first draft of the manuscript. SS and AM lead the contact tracing process. RD, AH, LW, and CC conceived of the project and designed the analytical approach. All authors edited the manuscript.

## Acknowledgements

The authors would like to acknowledge the substantial efforts of Newfoundland and Labrador Public Health, the Department of Health and Community Services, Regional Health Authorities and all individuals involved in contact tracing and the recording of these data, as well as the Newfoundland and Labrador Health Services, Digital Health as the custodians of these data.

